# COVID-19 among patients with hepatitis B or hepatitis C: A systematic review

**DOI:** 10.1101/2020.10.22.20216317

**Authors:** Hossein Mirzaie, Mohammad Vahidi, Mostafa Shokoohi, Maryam Darvishian, Hamid Sharifi, Heidar Sharafi, Mohammad Karamouzian

**Affiliations:** HIV/STI Surveillance Research Center, and WHO Collaborating Center for HIV Surveillance, Institute for Futures Studies in Health, Kerman University of Medical Sciences, Kerman, Iran; Faculty of Medicine, Shahid Beheshti University of Medical Sciences, Tehran, Iran; Division of Social and Behavioural Health Sciences, Dalla Lana School of Public Health, University of Toronto, Toronto, ON, Canada; Cancer Control Research, BC Cancer Research Centre, Vancouver, BC, Canada; Middle East Liver Diseases (MELD) Center, Tehran, Iran; School of Population and Public Health, Faculty of Medicine, University of British Columbia, Vancouver, BC, Canada

**Keywords:** COVID-19, Hepatitis B, Hepatitis C, Systematic Review

## Abstract

**Background & aims:** Hepatic manifestations of coronavirus disease 2019 (COVID-19) are common among people infected with hepatitis B virus (HBV) and hepatitis C virus (HCV). This systematic review aimed to summarize the evidence on COVID-19 patients with HBV or HCV co-infections.

**Methods:** We searched multiple electronic databases and preprint servers from December 1, 2019 to August 9, 2020. Studies were included if they reported quantitative empirical data on COVID-19 patients with HBV or HCV co-infections. Descriptive analyses were reported and data were narratively synthesized. Quality assessments was completed using the Joanna Briggs Institute critical appraisal tools.

**Results:** Out of the 941 identified records, 28 studies were included. Of the eligible studies, 235 patients with COVID-19 were infected with HBV and 22 patients with HCV. Most patients were male and mean age was 49.8 and 62.8 in patients with HBV and HCV, respectively. Death proportion was 6% among COVID-19-HBV and 13% among COVID-19-HCV co-infected patients. Among COVID-19 patients, 34.1% and 76.2% reported at least one comorbidity besides HBV and HCV infections, mainly hypertension and diabetes mellites type 2. The most common COVID-19-related symptoms in both HBV and HCV groups were fever, cough and dyspnea. ICU admission was reported in 14.1% and 21.4% of individuals with HBV and HCV, respectively.

**Conclusions:** Our findings suggest a considerable risk of morbidity and mortality among COVID-19 patients with HBV and HCV. Careful assessment of hepatic manifestations upon admission of patients could help improve health outcomes among COVID-19 patients with HBV or HCV co-infections.

**Key Points:**

- Hepatic manifestations of COVID-19 are common among people infected with HBV and HCV.
- Among COVID-19 patients, 34.1% and 76.2% reported at least one comorbidity besides HBV and HCV infections.
- The most common COVID-19-related symptoms in both HBV and HCV groups were fever, cough and dyspnea.
- There is a considerable risk of mortality among COVID-19 patients with HBV and HCV.

## INTRODUCTION

The first case of pneumonia caused by severe acute respiratory syndrome coronavirus 2 (SARS-CoV-2) was diagnosed in china in December 2019 (1). The world health organization (WHO) declared coronavirus disease 2019 (COVID-19) a pandemic on March 11, 2020. As of October 19, 2020, near the 40 million cases and more than 1,110,000 deaths had been reported worldwide (2). A more severe disease course and a higher mortality rate have been reported in older patients, as well as those with underlying medical conditions including hypertension, asthma, diabetes mellitus, chronic lung disease, cardiovascular conditions, obesity and chronic kidney disease (3-8). COVID-19 clinical presentations could vary greatly but often include respiratory, gastrointestinal, renal, and neurological manifestations (9, 10).

Recent studies have also reported COVID-19 cases presenting with hepatitis symptoms before developing respiratory symptoms (11). While our understanding of SARS-CoV-2’s pathogenesis continues to grow, initial studies suggest that the virus could lead to liver injury mainly by binding to angiotensin-converting enzyme 2 (ACE2) receptors on hepatocytes or causing an immune-mediated hepatic injury through cytokine storm activation (12-14). Several studies suggest that COVID-19 could lead to liver injuries and elevated alanine aminotransferase (ALT), aspartate aminotransferase (AST), and total bilirubin, particularly among sever COVID-19 cases (e.g., those admitted to intensive care unit [ICU](15-18). Abnormal liver functions among COVID-19 patients has also been associated with increased disease severity and risk of mortality (19, 20). A recent meta-analysis of a few studies on hepatic manifestations of COVID-19 estimated the pooled prevalence of pre-existing chronic liver disease as 1.9%, pre-existing liver cirrhosis as 0.4%, HBV as 0.9%, and HCV as 0.3% (15). While insightful, these findings are limited by small sample-sized studies conducted in China and USA during the early months of the pandemic (i.e., as late as April 2020) (15). These findings also contradict other earlier studies that reported a significantly higher prevalence of liver complications among COVID-19 patients in China where 2-11% of COVID-19 patients had liver-related complications and up to 54% had elevated AST and ALT levels (21).

COVID-19-related hepatic complications are particularly concerning among people living with HCV, or HBV, or HBV/HCV co-infection with pre-existing liver complications (e.g., cirrhosis, liver failure, hepatocellular carcinoma) (15, 21, 22). Considering that around 290 million and 71 million people are living with HBV and HCV, respectively (23, 24), the number of patients with SARS-CoV-2 and HBV and/or HCV co-infections are likely to increase. Therefore, improving our understanding of the hepatic manifestations and comorbidities among COVID-19 patients living with HBV, HCV, or both is of utmost importance in enhancing the care provided for this large at-risk population. In this systematic review, we aimed to review and summarize the existing literature on COVID-19 patients living with HBV or HCV infections. The findings of our review could provide information on clinical care and treatment options for COVID-19 patients with these infections.

## METHODS

Inclusion criteria and analytical plan were conceptualized a priori, and are documented in Open Science Framework (https://osf.io/p4w3j/).

### Literature search

Following the Systematic Reviews and Meta-Analyses (PRISMA) and Peer Review of Electronic Search Strategies (25) guidelines (see **supplementary file S1** for PRISMA checklist), we searched PubMed, Scopus, Web of Science, CINAHL, Embase, Google Scholar, as well as preprint databases including medRxiv and bioRxiv from December 1, 2019 to August 9, 2020. Search terms were combined using appropriate Boolean operators and included subject heading terms/keywords relevant to COVID-19 (e.g., SARS-CoV-2 or coronavirus disease 2019 or COVID-19 or severe acute respiratory syndrome coronavirus 2 or coronavirus infection) and hepatitis B or C (e.g., liver fibrosis or liver cirrhosis or hepatic transplantation or liver transplant or hepatitis C or hepatitis B or HBV or HCV). Please see **supplementary file S2** for our sample search strategy.

### Inclusion criteria and study selection

Quantitative studies of any type (i.e., case report, case series, cross-sectional, case–control, cohort, and clinical trial) that reported individual-level and/or aggregate-level data on COVID-19 patients living with HBV, HCV, or HBV-HCV-co-infection were included in this review. Studies that combined samples of HBV/HCV-positive and HBV/HCV-negative patients were only included if they provided subgroup analyses for people living with HBV, HCV or HBV-HCV-co-infection. Studies were excluded if they did not present original empirical data or did not report any clinical data for patients. Two reviewers (HM and MV) independently screened all titles, abstracts, and full-texts. Duplicate records were excluded and disagreements were resolved by discussion or arbitration by the senior author (MK).

### Data extraction

Data were extracted on a) *study characteristics*, including first author, publication date, study type, sample size, data type, and study population, b) *socio-demographic characteristics*, such as patients’ age and sex/gender, c) *HBV- and HCV-related characteristics*, including cirrhosis status, liver transplantation, receiving immunosuppressive therapy, liver enzyme levels (e.g., AST, ALT, ALP and GGT), d) *COVID-19-related characteristics*, including symptoms and severity of COVID-19 defined as mild (i.e., no or mild pneumonia), severe (i.e., blood oxygen saturation≤93%, dyspnea, or lung infiltrates >50% within two days], and critical (i.e., septic shock, respiratory failure, or multiple organ failures) (26). Hospitalization, ICU admission, survival status (recovery or death), as well as non-hepatic-related comorbidities (e.g., hypertension, diabetes, cardiovascular diseases, chronic obstructive pulmonary disease [COPD], and malignancies) were also recorded.

### Quality assessments

Two reviewers (HM and MV) completed quality assessments independently, using the Joanna Briggs Institute critical appraisal tools (27). These tools assess different items (e.g., selection bias, information bias, and confounding bias) for various study designs; nine items for case reports, ten for case series, nine for cross-sectional studies, and eleven for cohort studies.

### Statistical analysis

Descriptive analyses were used for reporting results. Continuous variables were summarized as mean and standard deviation (SD). Categorical variables were summarized by frequencies and percentages. Differences in continuous and categorical variables were compared using the two-tailed student’s t-test and Fisher’s exact test, respectively. P-values less than 0.05 were considered as statistically significant. For combining data from studies that reported aggregate-level data with those reporting individual-level data, we weighted the aggregate-level data by the number of patients. The proportion of death among reported patients was also measured and reported. The denominator and nominator for this measure were based on patients with HBV or HCV whose COVID-19 was diagnosed and reported. We also conducted a subgroup analysis by patients’ receipt of immunosuppressive medication.

## RESULTS

### Study characteristics

Out of 941 identified records, 28 studies met our inclusion criteria and were considered for data extraction. **Figure 1** shows the PRISMA flow diagram and study characteristics are presented in **Table 1**. Of the 28 included studies that reported some quantitative data on SARS-CoV-2 and HBV and/or HCV co-infections, 15 studies reported SARS-CoV-2 and HBV co-infection, 10 studies reported SARS-CoV-2 and HCV co-infection, and three reported both types of co-infections. With regards to their geographical location, 15 studies reported data from china, three from the US, two from Spain, two from Brazil, and the remaining six studies from Italy, Austria, Switzerland, the UK, Lithuania, and the UAE. All the included studies were observational: 11 studies were case reports, 10 were case series, and seven were cross-sectional. The majority of the included studies (n = 22) reported individual-level data, and six studies reported aggregate-level data.

**Table 1.**
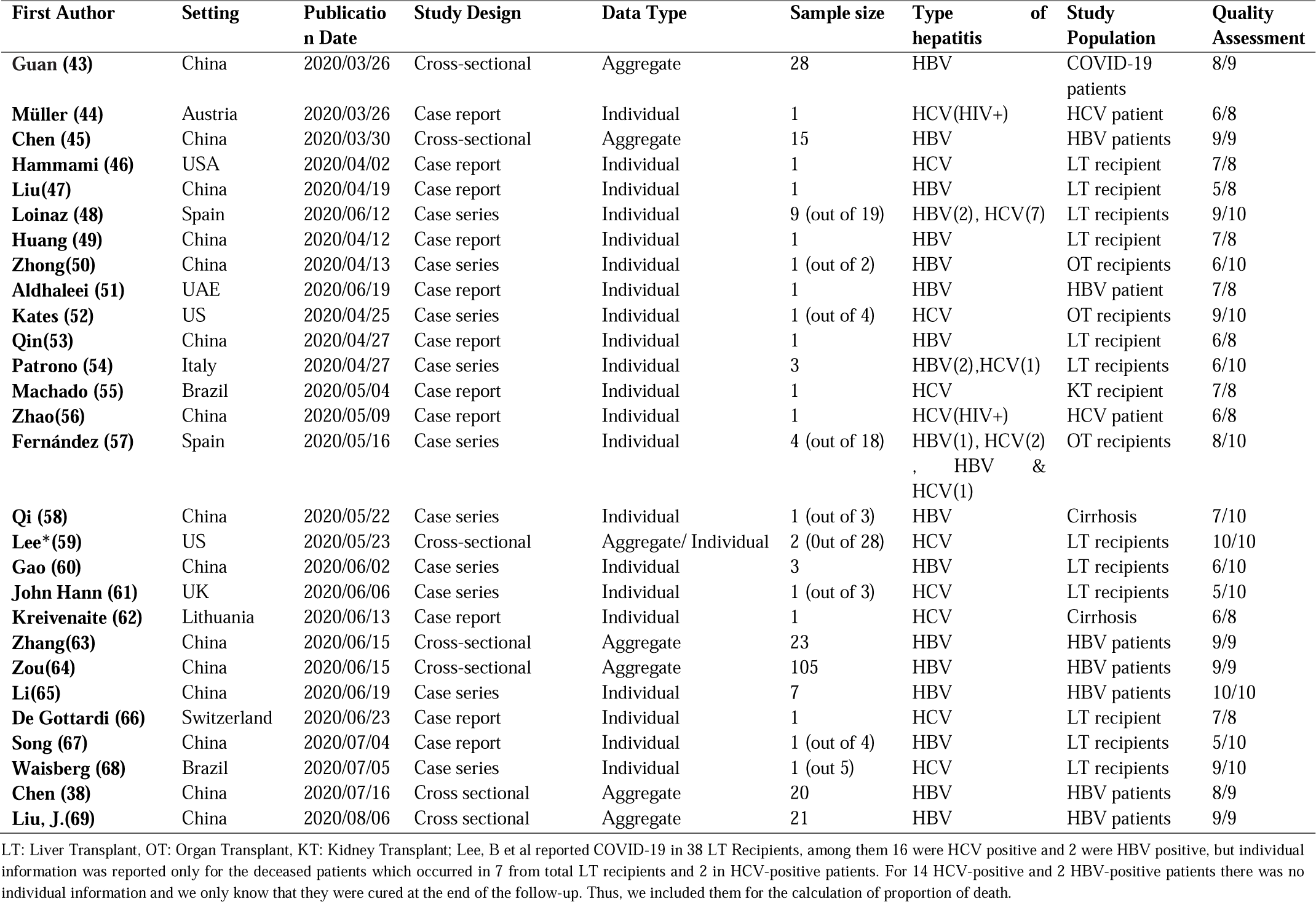
Characteristic of studies included in the review of SARS-CoV-2-HBV or SARS-CoV-2-HCV co-infections.

| First Author | Setting | Publication Date | Study Design | Data Type | Sample size | Type of hepatitis | Study Population | Quality Assessment |
| --- | --- | --- | --- | --- | --- | --- | --- | --- |
| Guan (43) | China | 2020/03/26 | Cross-sectional | Aggregate | 28 | HBV | COVID-19 patients | 8/9 |
| Müller (44) | Austria | 2020/03/26 | Case report | Individual | 1 | HCV(HIV+) | HCV patient | 6/8 |
| Chen (45) | China | 2020/03/30 | Cross-sectional | Aggregate | 15 | HBV | HBV patients | 9/9 |
| Hammami (46) | USA | 2020/04/02 | Case report | Individual | 1 | HCV | LT recipient | 7/8 |
| Liu(47) | China | 2020/04/19 | Case report | Individual | 1 | HBV | LT recipient | 5/8 |
| Loinaz (48) | Spain | 2020/06/12 | Case series | Individual | 9 (out of 19) | HBV(2), HCV(7) | LT recipients | 9/10 |
| Huang (49) | China | 2020/04/12 | Case report | Individual | 1 | HBV | LT recipient | 7/8 |
| Zhong(50) | China | 2020/04/13 | Case series | Individual | 1 (out of 2) | HBV | OT recipients | 6/10 |
| Aldhaleei (51) | UAE | 2020/06/19 | Case report | Individual | 1 | HBV | HBV patient | 7/8 |
| Kates (52) | US | 2020/04/25 | Case series | Individual | 1 (out of 4) | HCV | OT recipients | 9/10 |
| Qin(53) | China | 2020/04/27 | Case report | Individual | 1 | HBV | LT recipient | 6/8 |
| Patrono (54) | Italy | 2020/04/27 | Case series | Individual | 3 | HBV(2),HCV(1) | LT recipients | 6/10 |
| Machado (55) | Brazil | 2020/05/04 | Case report | Individual | 1 | HCV | KT recipient | 7/8 |
| Zhao(56) | China | 2020/05/09 | Case report | Individual | 1 | HCV(HIV+) | HCV patient | 6/8 |
| Fernández (57) | Spain | 2020/05/16 | Case series | Individual | 4 (out of 18) | HBV(1), HCV(2), HBV & HCV(1) | OT recipients | 8/10 |
| Qi (58) | China | 2020/05/22 | Case series | Individual | 1 (out of 3) | HBV | Cirrhosis | 7/10 |
| Lee*(59) | US | 2020/05/23 | Cross-sectional | Aggregate/ Individual | 2 (Out of 28) | HCV | LT recipients | 10/10 |
| Gao (60) | China | 2020/06/02 | Case series | Individual | 3 | HBV | LT recipients | 6/10 |
| John Hann (61) | UK | 2020/06/06 | Case series | Individual | 1 (out of 3) | HCV | LT recipients | 5/10 |
| Kreivenaite (62) | Lithuania | 2020/06/13 | Case report | Individual | 1 | HCV | Cirrhosis | 6/8 |
| Zhang(63) | China | 2020/06/15 | Cross-sectional | Aggregate | 23 | HBV | HBV patients | 9/9 |
| Zou(64) | China | 2020/06/15 | Cross-sectional | Aggregate | 105 | HBV | HBV patients | 9/9 |
| Li(65) | China | 2020/06/19 | Case series | Individual | 7 | HBV | HBV patients | 10/10 |
| De Gottardi (66) | Switzerland | 2020/06/23 | Case report | Individual | 1 | HCV | LT recipient | 7/8 |
| Song (67) | China | 2020/07/04 | Case report | Individual | 1 (out of 4) | HBV | LT recipients | 5/10 |
| Waisberg (68) | Brazil | 2020/07/05 | Case series | Individual | 1 (out 5) | HCV | LT recipients | 9/10 |
| Chen (38) | China | 2020/07/16 | Cross sectional | Aggregate | 20 | HBV | HBV patients | 8/9 |
| Liu, J.(69) | China | 2020/08/06 | Cross sectional | Aggregate | 21 | HBV | HBV patients | 9/9 |

LT: Liver Transplant, OT: Organ Transplant, KT: Kidney Transplant; Lee, B et al reported COVID-19 in 38 LT Recipients, among them 16 were HCV positive and 2 were HBV positive, but individual information was reported only for the deceased patients which occurred in 7 from total LT recipients and 2 in HCV-positive patients. For 14 HCV-positive and 2 HBV-positive patients there was no individual information and we only know that they were cured at the end of the follow-up. Thus, we included them for the calculation of proportion of death.

**Figure 1.**
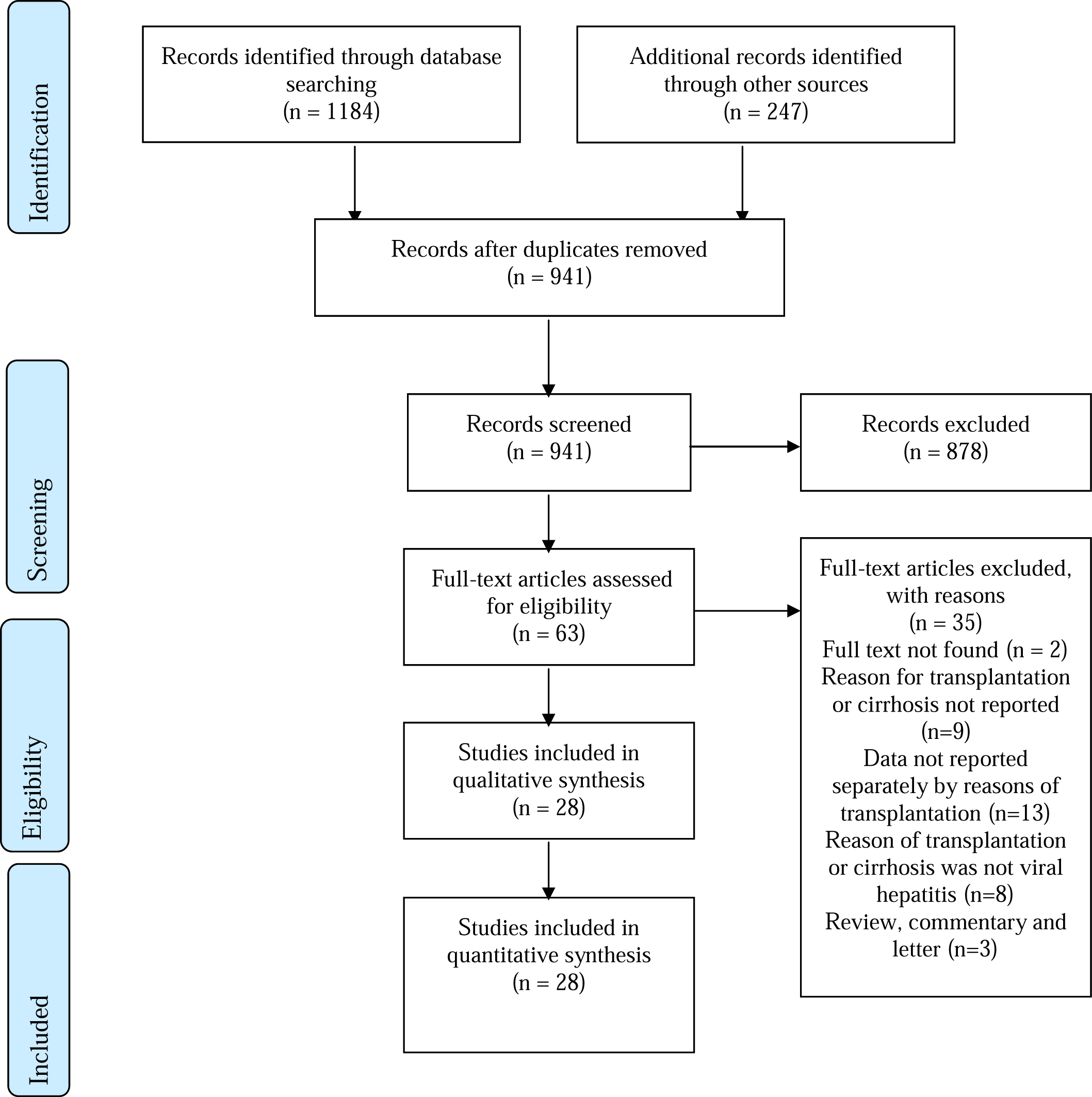
Flowchart of studies included in the systematic review of SARS-CoV-2-HBV or SARS-CoV-2-HCV co-infections

### Participant characteristics

As shown in **Table 1**, study populations were diverse and included organ transplant patients (16 studies), people living with HBV and/or HCV (nine studies), cirrhosis patients (two studies), and a diverse group of COVID-19 patients with hepatic co-infections (one study). Publication dates ranged from March 26, 2020 to August, 06, 2020. Across the 28 studies, 235 SARS-CoV-2 infections were found in patients with HBV and 22 in patients with HCV. Mean (SD) age was 49.8 (9.7) and 62.8 (10.3) among patients with HBV and HCV, respectively. Most patients in both groups were male (62.1% among patients with HBV and 72.8% among patients with HCV). Overall, other than transplanted patients, 10.2% (19 out of 186) of patients with HBV and 95.2% (21 out of 22) of patients with HCV were cirrhotic. A history of liver transplantation was found in 7.0% (13 out of 186) of patients with HBV and 90.9% (20 out of 22) of patients with HCV. Among patients with SARS-CoV-2 and HBV, 34.1% (61 out of 179) reported at least one comorbidity other than liver disease; most common comorbidities were hypertension (19.6%, 35 out of 179), diabetes mellitus type 2 (8.9%, 16 out of 179), cardiovascular diseases (4.5%, 8 out of 179) and any type of malignancy (4.5%, 8 out of 179). Among patients with SARS-CoV-2 and HCV, 76.2% (16 out of 21) reported at least one comorbidity other than liver disease; most common comorbidities were hypertension (57.1%, 12 out of 21) and diabetes mellitus type 2 (38.1%, 8 out of 21). see **Table 2** for further details.

**Table 2.** Demographic and clinical characteristics of COVID-19 patients with hepatitis B (n=235) and hepatitis C (n=22) included in the reviewed studies

| Characteristics (n reported)* | HBV<br>N (%) | HCV<br>N (%) |
| --- | --- | --- |
| **Age (Mean ± SD)** | 49.8 ± 9.7 [n=110] | 62.8±10.3 [n=22] |
| **Sex** | n=235 | n=22 |
| Male | 146 (62.1) | 16 (72.8) |
| Female | 89 (37.9) | 6 (27.3) |
| **Cirrhosis** | n=186 | n=21 |
| Yes | 19 (10.2) | 20 (95.2) |
| **Liver transplantation** | n=186 | N=22 |
| Yes | 13 (7.0) | 20 (90.9) |
| **Immunosuppressive therapy** | N=186 | N=22 |
| Yes | 13 (7.0) | 20 (90.9) |
| **Elevated liver biochemistries** | N=163 | N= 13 |
| ALT | 40 (24.5) | 5 (38.5) |
| AST | 43 (26.4) | 5 (38.5) |
| ALP | 4 (1.8) | 3 (23.1) |
| GGT | 24 (10.4) | 3 of 9 (33.3) |
| **Co-morbidities (in addition to COVID-19/Hepatitis)** | n=179 | n=21 |
| At least one morbidity | 61 (34.1) | 16 (76.2) |
| Hypertension | 35 (19.6) | 12 (57.1) |
| Chronic obstructive pulmonary disease | 5 (2.8) | 1 (4.8) |
| Diabetes | 16 (8.9) | 8 (38.1) |
| Cardiovascular disease | 8 (4.5) | 4 (19) |
| Malignancy | 8 (4.5) | 0 |
| **COVID-19 symptoms** | n=191 | n= 22 |
| Fever | 150 (78.5) | 17 (77.3) |
| Cough | 112 (58.6) | 14 (63.6) |
| Dyspnea | 74 (38.7) | 9 (40.9) |
| Headache | 5 (2.6) | 1 (4.5) |
| Arthralgia/myalgia | 8 (4.2) | 4 (18.2) |
| Fatigue | 60 (31.4) | 4 (18.2) |
| Gastrointestinal symptoms | 28 (14.6) | 7 (31.8) |
| **Severity\*** | n=233 | N= 14 |
| Non-Severe | 143 (61.4) | 11 (78.6) |
| Severe | 90 (38.6) | 3 (21.4) |
| **Hospitalized** | n=234 | n=22 |
| Yes | 234 (100) | 19 (86.4) |
| **Intensive care unit admission** | n=92 | N=14 |
| Yes | 13 (14.1) | 3 (21.4) |
| **Death** | n=234 | N=22 |
| Yes | 14 (6.0) | 3 (13.6) |

\*Mild cases reported as non-severe and severe/critical cases are reported as severe.

### COVID-19-related symptoms

As reported in **Table 2**, the most common COVID-19-related symptoms in patients with SARS-CoV-2 and HBV were fever (78.5%, 150 out of 191), cough (58.6%, 112 out of 191), dyspnea (38.7%, 74 out of 191), fatigue (31.4%, 60 out of 191), and gastrointestinal symptoms (14.6%, 28 out of 191). Most common symptoms in patients with SARS-CoV-2 and HCV were fever (77.3%, 17 out of 22), cough (63.6%, 14 out of 22), dyspnea (40.9%, 9 out of 22), and gastrointestinal symptoms (31.8%, 7 out of 22). Having elevated alanine aminotransferase (i.e., ALT >45 IU/L) was found in 24.5% (40 out of 163 patients) in patients with SARS-CoV-2 and HBV and 38.5% (5 of 13) in patients with SARS-CoV-2 and HCV. In addition, elevated aspartate aminotransferase (i.e., AST >45 IU/L or AST >45 IU/L) was found in 26.5% (43 of 163 patients) in SARS-CoV-2-HBV group and 38.5% (5 of 13) in SARS-CoV-2-HCV group. All of the patients in the SARS-CoV-2-HBV infected group, and 86.4% (19 of 22) case in SARS-CoV-2-HCV infected group were hospitalized. ICU admission was reported in 14.1% in the HBV group and 21.4% in the HCV group. Severe COVID-19 was reported in 38.6% (90 of 233) in HBV patients and 21.4% (3 of 14) in HCV patients.

### Death proportion among the patients

Of all 235 reported cases of SARS-CoV-2-HBV co-infected patients, 6% (14 cases) were deceased. Information on the age of the deceased patients in this group was available for 6 of 14 patients. The mean (SD) age of deceased patients was 59.3 (13.4) years, which was higher than the mean (SD) age of all included patients estimated as 49.8 (9.7). Information on the sex of the deceased patients was available for 6 (out of 14) patients, and all of them were male. Three of 14 deceased patients were taking immunosuppressive therapy due to liver transplantation. Of all 22 reported cases of SARS-CoV-2-HCV coinfected patients, 13.6% (three people) passed away. These three cases were 69, 71, and 79 years old. Two of them were female, and one was male. They had more than one comorbidity and took immunosuppressive therapy due to liver transplantation.

### COVID-19 among immunosuppressed patients

Based on available individual data, the mean (SD) age of immunosuppressed patients was significantly higher than patients without immunosuppression (60.8 (11.7) vs 49.9 (12); P-value = 0.001). 85.3% (n = 29) of immunosuppressed patients and 66% (n = 31) of patients without immunosuppression ware male (P-value = 0.04). 51.5% (n = 17) of patients with immunosuppression and 30% (n = 12) of patients without immunosuppression have had at least one comorbidity other than COVID-19 and hepatitis (P-value = 0.07). Gastrointestinal symptoms were more common in immunosuppressed patients (32.2% vs 7.3%; P-value = 0.03). The proportion of severe COVID-19 in patients with and without immunosuppression was 33.3% and 30.4%, respectively (P-value = 0.7). 29.2% of patients with immunosuppression and 12.5% of patients without immunosuppression were admitted to the intensive care unit (P-value = 0.1). Death happened in 17.6% of patients with immunosuppression and 6.4% of patients without immunosuppression (P-value = 0.1). Please see **Table 3** for further details.

**Table 3.** Demographic and clinical characteristics of COVID-19 infection in hepatitis B and hepatitis C patients included in the reviewed studies, stratified by immunosuppression (IS) status

| **Characteristics (n reported)\*** | **IS therapy**<br>**N (%)** | **No IS therapy**<br>**N (%)** | **P-value** |
| --- | --- | --- | --- |
| **Age (Mean ± SD)** | 60.8 ± 11.7 | (n=22) 49.9±12 | 0.001 |
| **Sex** | n=34 | n=47 |  |
| Male | 29 (85.3) | 31 (66) | 0.04 |
| Female | 5 (14.7) | 16 (34) |  |
| **Co-morbidities (in addition to COVID-19 and hepatitis)** | n=33 | n=40 |  |
| At least one morbidity | 17 (51.5) | 12 (30) | 0.09 |
| **COVID-19 symptoms** | n=34 | n= 47 |  |
| Fever | 27 (79.4) | 29 (61.7) | 0.1 |
| Cough | 20 (58.8) | 23 (48.9) | 0.3 |
| Dyspnea | 13 (38.2) | 9 (19.1) | 0.09 |
| Headache | 1 (2.9) | 2 (4.2) | 0.3 |
| Arthralgia/myalgia | 6 (17.6) | 4 (8.5) | 0.2 |
| Fatigue | 6 (17.6) | 9 (19.1) | 0.1 |
| Gastrointestinal symptoms | 11 (32.3) | 3 (7.3) | 0.03 |
| **Severity** | n=24 | n= 46 |  |
| Non-Severe | 16 (66.7) | 32 (69.6) | 0.7 |
| Severe | 8 (33.3) | 14 (30.4) |  |
| **Hospitalized** | n=34 | n=47 | 0.07 |
| Yes | 31 (91.2) | 47 (100) |  |
| **Intensive care unit admission** | n=24 | n=32 | 0.1 |
| Yes | 7 (29.2) | 4 (12.5) |  |
| **Death** | n=34 | n=47 | 0.1 |
| Yes | 6 (17.6) | 3 (6.4) |  |

### Quality assessment

As shown in **Table 1**, quality assessment scores of the studies ranged from 5 to 7 for case reports (out of 8 possible points), 5 to 10 for the case series (out of 10 possible points) and 8 to 9 for cross-sectional (out of 9 possible points). Further details on quality assessment tools and respective scores are presented in **supplementary file S3**.

## DISCUSSION

We summarized data from 235 patients with SARS-CoV-2 and HBV co-infection and 22 patients with SARS-CoV-2 and HCV co-infection. While our findings are relatively comparable with evidence from COVID-19 patients without hepatic comorbidities (10, 28), our review pointed to higher morbidity and mortality among COVID-19 patients living with HBV, HCV, or both. The most common COVID-19-related symptoms were fever, cough, dyspnea, fatigue, and gastrointestinal symptoms which have also been common in COVID-19 patients without these co-infections (13, 29-31). Moreover, about 24% of patients with SARS-CoV-2-HBV co-infection and 39% of patients with SARS-CoV-2-HCV coinfection had elevated levels of ALT and AST which is in line with findings of a previous systematic review of earlier studies on liver manifestations among COVID-19 patients, where the pooled incidence of elevated liver chemistries among COVID-19 patients was estimated as 23.1% (32).

Many studies have reported elevated liver biochemistries to be common among COVID-19 patients with and without hepatic comorbidities (32-34). However, these findings should be interpreted with caution. First, it is unclear whether the liver damage observed among COVID-19 patients with HBV and HCV co-infections is mainly due to the adverse impact of SARS-CoV-2 on hepatic cells or patients’ pre□existing viral hepatitis. Second, corticosteroids that are commonly used as a treatment strategy in COVID-19 patients have been shown to increase the risk of hepatitis flare in HBV patients and might impact liver enzyme profiles among these patients (35-37). On the other hand, Chen et al. reported no difference in the level of liver function parameters between COVID-19 infected patients and COVID-19-HBV coinfected patients (38) and Kulkarni et al.’s meta-analysis of COVID-19-related liver manifestations reported patients with pre-existing chronic liver to have lower odds of developing severe COVID□19(32). All of these rather equivocal findings highlight the importance of conducting well-controlled studies to fully understand the pathogenesis of SARS-CoV-2 among HBV and HCV patients and determine the main underlying causes of elevated liver enzymes among these multimorbid patients to help inform clinical decision making.

All of the patients in the SARS-CoV-2-HBV group, and 86.4% (19 of 22) case in SARS-CoV-2-HCV group were hospitalized. While this finding might be interpreted as higher risk of hospitalization among these multimorbid population, it could also be reflective of the fact that the existing evidence is mainly skewed towards hospitalized sever COVID-19 patients with hepatic comorbidities. This was further evident in our finding that 14.1% in the HBV group and 21.4% in the HCV group were admitted to ICU and sever COVID-19 was reported in 38.8% of HBV patients and 21.4% of HCV patients. Moreover, 6% of the reported cases of SARS-CoV-2-HBV coinfection, and 13.6% of reported cases of SARS-CoV-2-HCV coinfection, were deceased. Given the high prevalence of multimorbidities among HBV and HCV patients and the fact that most patients included in this review were mostly sever cases of hospitalized COVID-19 with a history of organ transplant, this finding should also be interpreted with caution when compared to the estimated 2% overall risk of mortality among COVID-19 patients (10). Nonetheless, these findings are informative and suggest that among those who died, multimorbidity, older age, and male gender were common in comparison with all included patients.

Our findings on the impact of immunosuppression on COVID-19’s severity are interesting, given the equivocal findings of different studies on the susceptibility of immunosuppressed patients to severe SARS-CoV-2 infection. While some studies assume a higher risk for immunosuppressed population due to their systemic immunocompromised status (39, 40), others suggest immunosuppression may provide protection against hyperactivated immune response (41, 42). We found that the proportion of severe COVID-19 cases, admission to ICU, and proportion of death among patients with immunosuppression were higher than patients without immunosuppression; however, these differences were not statistically significant. These findings are not conclusive and should be interpreted considering the small sample size, the observational nature, the non-matched nature of the comparisons in the included studies, and the fact that these differences can be due to the immunosuppressed group’s higher likelihood for being older, male, and having had organ transplants, advanced liver disease, and other extrahepatic comorbidities.

We acknowledge the limitations of our study. First, while most COVID-19 patients are asymptomatic, the available evidence which informed this review included mostly hospitalized patients and is skewed towards more severe patients often with a history of organ transplant and advanced liver disease; therefore, our findings are not generalizable to all patients living with COVID-19 and HBV or HCV co-morbidities. Second due to descriptive design of included studies and lack of comparison group we could not identify factors associated with SARS-CoV-2-HBV or HCV co-infections. Third, most of the included studies had a small sample size and without a population-based survey of patients with HBV or HCV, the accurate prevalence of COVID-19 and its manifestations among HBV or HCV subpopulations remain unknown. Lastly, given the growing nature of the pandemic and the overwhelmed healthcare systems worldwide, viral hepatitis manifestations may be under-recorded or overlooked during clinical visits and therefore, underestimate the scope of hepatic manifestations among COVID-19 with HBV and HCV co-infections.

### Conclusion

Our findings suggest that COVID-19 patients with HBV and HCV co-infections may be at an increased risk of morbidity and mortality. Despite the limitations of the existing evidence, our review suggests that liver enzyme abnormalities and acute hepatic injuries may be common among COVID-19 patients with HBV and HCV co-infections. Therefore, these paraclinical profiles should be monitored and examined during clinical visits. While understanding the pathogenesis of SARS-CoV-2 requires further investigations, careful assessment of hepatic manifestations upon admission could help reduce the multimorbidity among HBV or HCV patients and lead to more favourable health outcomes among COVID-19 patients.

## Data Availability

All data is presented in the paper

## Acknowledgements

The authors did not receive any funding for this study. MK is a member of Pierre Elliott Trudeau Foundation’s COVID-19 impact committee and is supported by the Vanier Canada Graduate Scholarship and the Pierre Elliott Trudeau Foundation Doctoral Scholarship. MS is supported by a Canadian Institutes of Health Research (CIHR) Postdoctoral Scholarship.

## REFERENCES

1. Zhu N, Zhang D, Wang W, Li X, Yang B, Song J, et al. A Novel Coronavirus from Patients with Pneumonia in China, 2019. N Engl J Med. 2020;382(8):727–33.

2. Organization WH. WHO coronavirus disease (COVID-19) dashboard: WHO; 2020 [19 October 2020]. Available from: https://covid19.who.int/.

3. Wang L, He W, Yu X, Hu D, Bao M, Liu H, et al. Coronavirus disease 2019 in elderly patients: Characteristics and prognostic factors based on 4-week follow-up. J Infect. 2020;80(6):639–45.

4. Guo W, Li M, Dong Y, Zhou H, Zhang Z, Tian C, et al. Diabetes is a risk factor for the progression and prognosis of COVID-19. Diabetes Metab Res Rev. 2020:e3319.

5. Mostaza JM, García-Iglesias F, González-Alegre T, Blanco F, Varas M, Hernández-Blanco C, et al. Clinical course and prognostic factors of COVID-19 infection in an elderly hospitalized population. Arch Gerontol Geriatr. 2020;91:104204.

6. Földi M, Farkas N, Kiss S, Zádori N, Váncsa S, Szakó L, et al. Obesity is a risk factor for developing critical condition in COVID-19 patients: A systematic review and meta-analysis. Obes Rev. 2020;21(10):e13095.

7. Fang X, Li S, Yu H, Wang P, Zhang Y, Chen Z, et al. Epidemiological, comorbidity factors with severity and prognosis of COVID-19: a systematic review and meta-analysis. Aging (Albany NY). 2020;12(13):12493–503.

8. Figliozzi S, Masci PG, Ahmadi N, Tondi L, Koutli E, Aimo A, et al. Predictors of adverse prognosis in COVID-19: A systematic review and meta-analysis. Eur J Clin Invest. 2020;50(10):e13362.

9. Goyal P, Choi JJ, Pinheiro LC, Schenck EJ, Chen R, Jabri A, et al. Clinical Characteristics of Covid-19 in New York City. N Engl J Med. 2020;382(24):2372–4.

10. Khalili M, Karamouzian M, Nasiri N, Javadi S, Mirzazadeh A, Sharifi H. Epidemiological characteristics of COVID-19: a systematic review and meta-analysis. Epidemiol Infect. 2020;148:e130.

11. Wander P, Epstein M, Bernstein D. COVID-19 Presenting as Acute Hepatitis. Am J Gastroenterol. 2020;115(6):941–2.

12. Alqahtani SA, Schattenberg JM. Liver injury in COVID-19: The current evidence. United European Gastroenterol J. 2020;8(5):509–19.

13. Kulkarni AV, Kumar P, Tevethia HV, Premkumar M, Arab JP, Candia R, et al. Systematic review with meta-analysis: liver manifestations and outcomes in COVID-19. Aliment Pharmacol Ther. 2020;52(4):584–99.

14. Mehta P, McAuley DF, Brown M, Sanchez E, Tattersall RS, Manson JJ. COVID-19: consider cytokine storm syndromes and immunosuppression. Lancet. 2020;395(10229):1033–4.

15. Kunutsor SK, Laukkanen JA. Hepatic manifestations and complications of COVID-19: A systematic review and meta-analysis. J Infect. 2020;81(3):e72–e4.

16. Huang C, Wang Y, Li X, Ren L, Zhao J, Hu Y, et al. Clinical features of patients infected with 2019 novel coronavirus in Wuhan, China. Lancet. 2020;395(10223):497–506.

17. Guan WJ, Ni ZY, Hu Y, Liang WH, Ou CQ, He JX, et al. Clinical Characteristics of Coronavirus Disease 2019 in China. N Engl J Med. 2020;382(18):1708–20.

18. Khinda J, Janjua NZ, Cheng S, van den Heuvel ER, Bhatti P, Darvishian M. Association between markers of immune response at hospital admission and COVID-19 disease severity and mortality: A meta-analysis and meta-regression. Journal of medical virology. 2020.

19. Fan Z, Chen L, Li J, Cheng X, Yang J, Tian C, et al. Clinical Features of COVID-19-Related Liver Functional Abnormality. Clinical gastroenterology and hepatology : the official clinical practice journal of the American Gastroenterological Association. 2020;18(7):1561–6.

20. Cai Q, Huang D, Yu H, Zhu Z, Xia Z, Su Y, et al. COVID-19: Abnormal liver function tests. J Hepatol. 2020;73(3):566–74.

21. Zhang C, Shi L, Wang FS. Liver injury in COVID-19: management and challenges. Lancet Gastroenterol Hepatol. 2020;5(5):428–30.

22. Hernandez MDP, Martin P, Simkins J. Infectious complications after liver transplantation. Gastroenterology & hepatology. 2015;11(11):741.

23. Razavi-Shearer D, Gamkrelidze I, Nguyen MH, Chen D-S, Van Damme P, Abbas Z, et al. Global prevalence, treatment, and prevention of hepatitis B virus infection in 2016: a modelling study. The lancet Gastroenterology & hepatology. 2018;3(6):383–403.

24. Blach S, Zeuzem S, Manns M, Altraif I, Duberg A-S, Muljono DH, et al. Global prevalence and genotype distribution of hepatitis C virus infection in 2015: a modelling study. The lancet Gastroenterology & hepatology. 2017;2(3):161–76.

25. Pressman P, Clemens R, Sahu S, Hayes AW. A Review of Methanol Poisoning: A Crisis Beyond Ocular Toxicology. Cutaneous and ocular toxicology. 2020:1-19.

26. Mirzaei H, McFarland W, Karamouzian M, Sharifi H. COVID-19 Among People Living with HIV: A Systematic Review. AIDS Behav. 2020:1-8.

27. Joanna Briggs Institute. JBI’s Critical Appraisal Tools 2020 [Available from: https://joannabriggs.org/critical-appraisal-tools.

28. Rodriguez-Morales AJ, Cardona-Ospina JA, Gutiérrez-Ocampo E, Villamizar-Peña R, Holguin-Rivera Y, Escalera-Antezana JP, et al. Clinical, laboratory and imaging features of COVID-19: A systematic review and meta-analysis. Travel medicine and infectious disease. 2020;34:101623.

29. Hu Y, Sun J, Dai Z, Deng H, Li X, Huang Q, et al. Prevalence and severity of corona virus disease 2019 (COVID-19): A systematic review and meta-analysis. Journal of clinical virology : the official publication of the Pan American Society for Clinical Virology. 2020;127:104371.

30. Chen N, Zhou M, Dong X, Qu J, Gong F, Han Y, et al. Epidemiological and clinical characteristics of 99 cases of 2019 novel coronavirus pneumonia in Wuhan, China: a descriptive study. Lancet. 2020;395(10223):507–13.

31. Zhu J, Ji P, Pang J, Zhong Z, Li H, He C, et al. Clinical characteristics of 3062 COVID-19 patients: A meta-analysis. Journal of medical virology. 2020:10.1002/jmv.25884.

32. Kulkarni AV, Kumar P, Tevethia HV, Premkumar M, Arab JP, Candia R, et al. Systematic review with meta-analysis: liver manifestations and outcomes in COVID-19. Alimentary pharmacology & therapeutics. 2020;52(4):584–99.

33. Yang X, Yu Y, Xu J, Shu H, Xia Ja, Liu H, et al. Clinical course and outcomes of critically ill patients with SARS-CoV-2 pneumonia in Wuhan, China: a single-centered, retrospective, observational study. Lancet Respir Med. 2020;8(5):475–81.

34. Xu X-W, Wu X-X, Jiang X-G, Xu K-J, Ying L-J, Ma C-L, et al. Clinical findings in a group of patients infected with the 2019 novel coronavirus (SARS-Cov-2) outside of Wuhan, China: retrospective case series. bmj. 2020;368.

35. Shang L, Zhao J, Hu Y, Du R, Cao B. On the use of corticosteroids for 2019-nCoV pneumonia. Lancet. 2020;395(10225):683–4.

36. Wong GL, Wong VW, Yuen BW, Tse YK, Yip TC, Luk HW, et al. Risk of hepatitis B surface antigen seroreversion after corticosteroid treatment in patients with previous hepatitis B virus exposure. J Hepatol. 2020;72(1):57–66.

37. Hwang JP, Lok AS. Management of patients with hepatitis B who require immunosuppressive therapy. Nat Rev Gastroenterol Hepatol. 2014;11(4):209–19.

38. Chen L, Huang S, Yang J, Cheng X, Shang Z, Lu H, et al. Clinical characteristics in patients with SARS-CoV-2/HBV co-infection. Journal of viral hepatitis. 2020:10.1111/jvh.13362.

39. Jin X, Lian J-S, Hu J-H, Gao J, Zheng L, Zhang Y-M, et al. Epidemiological, clinical and virological characteristics of 74 cases of coronavirus-infected disease 2019 (COVID-19) with gastrointestinal symptoms. Gut. 2020;69(6):1002–9.

40. Zhang C, Shi L, Wang F-S. Liver injury in COVID-19: management and challenges. The lancet Gastroenterology & hepatology. 2020;5(5):428–30.

41. D’Antiga L. Coronaviruses and Immunosuppressed Patients: The Facts During the Third Epidemic. Liver Transpl. 2020;26(6):832–4.

42. Boettler T, Newsome PN, Mondelli MU, Maticic M, Cordero E, Cornberg M, et al. Care of patients with liver disease during the COVID-19 pandemic: EASL-ESCMID position paper. JHEP Rep. 2020;2(3):100113.

43. Guan W-j, Liang W-h, Zhao Y, Liang H-r, Chen Z-s, Li Y-m, et al. Comorbidity and its impact on 1,590 patients with COVID-19 in China: A Nationwide Analysis. medRxiv. 2020:2020.02.25.20027664.

44. Müller H, Kniepeiss D, Stauber R, Schrem H, Rauter M, Krause R, et al. Recovery from COVID-19 following hepatitis C, human immunodeficiency virus infection, and liver transplantation. American journal of transplantation : official journal of the American Society of Transplantation and the American Society of Transplant Surgeons. 2020:10.1111/ajt.16107.

45. Chen X, Jiang Q, Ma Z, Ling J, Hu W, Cao Q, et al. Clinical Characteristics of Hospitalized Patients with SARS-CoV-2 and Hepatitis B virus Co-infection. medRxiv. 2020.

46. Hammami MB, Garibaldi B, Shah P, Liu G, Jain T, Chen PH, et al. Clinical course of COVID-19 in a liver transplant recipient on hemodialysis and response to tocilizumab therapy: A case report. American journal of transplantation : official journal of the American Society of Transplantation and the American Society of Transplant Surgeons. 2020;20(8):2254–9.

47. Liu B, Wang Y, Zhao Y, Shi H, Zeng F, Chen Z. Successful treatment of severe COVID-19 pneumonia in a liver transplant recipient. American journal of transplantation : official journal of the American Society of Transplantation and the American Society of Transplant Surgeons. 2020;20(7):1891–5.

48. Loinaz C, Marcacuzco A, Fernández-Ruiz M, Caso O, Cambra F, San Juan R, et al. Varied clinical presentation and outcome of SARS-CoV-2 infection in liver transplant recipients: Initial experience at a single center in Madrid, Spain. Transplant infectious disease : an official journal of the Transplantation Society. 2020:e13372.

49. Huang JF, Zheng KI, George J, Gao HN, Wei RN, Yan HD, et al. Fatal outcome in a liver transplant recipient with COVID-19. American journal of transplantation : official journal of the American Society of Transplantation and the American Society of Transplant Surgeons. 2020;20(7):1907–10.

50. Zhong Z, Zhang Q, Xia H, Wang A, Liang W, Zhou W, et al. Clinical characteristics and immunosuppressant management of coronavirus disease 2019 in solid organ transplant recipients. American journal of transplantation : official journal of the American Society of Transplantation and the American Society of Transplant Surgeons. 2020;20(7):1916–21.

51. Aldhaleei WA, Alnuaimi A, Bhagavathula AS. COVID-19 Induced Hepatitis B Virus Reactivation: A Novel Case From the United Arab Emirates. Cureus. 2020;12(6):e8645.

52. Kates OS, Fisher CE, Stankiewicz-Karita HC, Shepherd AK, Church EC, Kapnadak SG, et al. Earliest cases of coronavirus disease 2019 (COVID-19) identified in solid organ transplant recipients in the United States. American journal of transplantation : official journal of the American Society of Transplantation and the American Society of Transplant Surgeons. 2020;20(7):1885–90.

53. Qin J, Wang H, Qin X, Zhang P, Zhu L, Cai J, et al. Perioperative Presentation of COVID-19 Disease in a Liver Transplant Recipient. Hepatology (Baltimore, Md). 2020;72(4):1491–3.

54. Patrono D, Lupo F, Canta F, Mazza E, Mirabella S, Corcione S, et al. Outcome of COVID-19 in liver transplant recipients: A preliminary report from Northwestern Italy. Transplant infectious disease : an official journal of the Transplantation Society. 2020:e13353.

55. de Barros Machado DJ, Ianhez LE. COVID-19 pneumonia in kidney transplant recipients-where we are?Transplant Infectious Disease. 2020:e13306.

56. Zhao J, Liao X, Wang H, Wei L, Xing M, Liu L, et al. Early virus clearance and delayed antibody response in a case of COVID-19 with a history of co-infection with HIV-1 and HCV. Clinical infectious diseases : an official publication of the Infectious Diseases Society of America. 2020:ciaa408.

57. Fernández-Ruiz M, Andrés A, Loinaz C, Delgado JF, López-Medrano F, San Juan R, et al. COVID-19 in solid organ transplant recipients: A single-center case series from Spain. American Journal of Transplantation. 2020;20(7):1849–58.

58. Qi X, Wang J, Li X, Wang Z, Liu Y, Yang H, et al. Clinical course of COVID-19 in patients with pre-existing decompensated cirrhosis: initial report from China. Hepatology international. 2020;14(4):478–82.

59. Lee BT, Perumalswami PV, Im GY, Florman S, Schiano TD, Group CS. COVID-19 in Liver Transplant Recipients: An Initial Experience From the US Epicenter. Gastroenterology. 2020;159(3):1176-8.e2.

60. Zhen-Dong Y, Gao-Jun Z, Run-Ming J, Zhi-Sheng L, Zong-Qi D, Xiong X, et al. Clinical and transmission dynamics characteristics of 406 children with coronavirus disease 2019 in China: A review. The Journal of infection. 2020.

61. John Hann A, Lembach H, McKay SC, Perrin M, Isaac J, Oo YH, et al. Controversies regarding shielding and susceptibility to COVID-19 disease in liver transplant recipients in the United Kingdom. Transplant infectious disease : an official journal of the Transplantation Society. 2020:e13352.

62. Kreivenaite E, Gedgaudas R, Valantiene I, Mickiene A, Kupcinskas J. COVID-19 in a Patient with Liver Cirrhosis. Journal of gastrointestinal and liver diseases : JGLD. 2020;29(2):263–6.

63. Zhang B, Huang W, Zhang S. Clinical Features and Outcomes of Coronavirus Disease 2019 (COVID-19) Patients With Chronic Hepatitis B Virus Infection. Clinical Gastroenterology and Hepatology. 2020;18(11):2633–7.

64. Zou X, Fang M, Li S, Wu L, Gao B, Gao H, et al. Characteristics of Liver Function in Patients With SARS-CoV-2 and Chronic HBV Coinfection. Clinical gastroenterology and hepatology : the official clinical practice journal of the American Gastroenterological Association. 2020:S1542–3565(20)30821–1.

65. Li Y, Li C, Wang J, Zhu C, Zhu L, Ji F, et al. A case series of COVID-19 patients with chronic hepatitis B virus infection. Journal of medical virology. 2020:10.1002/jmv.26201.

66. De Gottardi A, Fratila C, Bertoli R, Cerny A, Magenta L, Gianella P, et al. Clinical characteristics and management of a liver transplanted patient admitted with SARS-CoV-2 infection. Clinics and research in hepatology and gastroenterology. 2020:S2210–7401(20)30159–5.

67. Song SH, Chen TL, Deng LP, Zhang YX, Mo PZ, Gao SC, et al. Clinical characteristics of four cancer patients with SARS-CoV-2 infection in Wuhan, China. Infectious diseases of poverty. 2020;9(1):82.

68. Waisberg DR, Abdala E, Nacif LS, Haddad LB, Ducatti L, Santos VR, et al. Liver transplant recipients infected with SARS-CoV-2 in the early postoperative period: Lessons from a single center in the epicenter of the pandemic. Transplant infectious disease : an official journal of the Transplantation Society. 2020:e13418.

69. Liu J, Wang T, Cai Q, Sun L, Huang D, Zhou G, et al. Longitudinal changes of liver function and hepatitis B reactivation in COVID-19 patients with pre-existing chronic hepatitis B virus infection. Hepatology research : the official journal of the Japan Society of Hepatology. 2020:10.1111/hepr.13553.

