## Supplement S2 for "COVID-19 among patients with hepatitis B or hepatitis C: A systematic review"

SUPPLEMENTARY FILE S2

**Sample Embase Search Strategy**

1. 'hepatitis b'/exp

2. 'acute hepatitis b'/exp

3. 'chronic hepatitis b'/exp

4. 'hepatitis b virus infection':ab,ti OR 'hepatitis, serum':ab,ti OR 'hippie hepatitis':ab,ti OR 'injection hepatitis':ab,ti OR 'serum hepatitis':ab,ti OR 'type b hepatitis':ab,ti OR 'viral hepatitis type b':ab,ti OR 'virus hepatitis type b':ab,ti OR 'hbv':ab,ti OR 'hep b':ab,ti OR 'hepatitis b':ab,ti

5. 'hepatitis c'/exp

6. 'acute hepatitis c'/exp

7. 'chronic hepatitis c'/exp

8. 'hepatitis c virus infection':ab,ti OR hcv:ab,ti OR 'hep c':ab,ti OR 'hepatitis c':ab,ti OR 'type c hepatitis':ab,ti OR 'viral hepatitis type c':ab,ti OR 'virus hepatitis type c':ab,ti

9. 'liver cirrhosis'/exp

10. 'liver cirrhosis':ab,ti

11. 'liver transplantation'/exp

12. 'auxiliary liver transplantation':ab,ti OR 'hepatic transplantation':ab,ti OR 'liver heterotopic transplantation':ab,ti OR 'liver orthotopic transplantation':ab,ti OR 'liver tissue transplantation':ab,ti OR 'orthotopic liver transplantation':ab,ti OR 'liver transplantation':ab,ti OR 'transplantation, liver':ab,ti

13. #1 OR #2 OR #3 OR #4 OR #5 OR #6 OR #7 OR #8 OR #9 OR #10 OR #11 OR #12

14. 'coronavirus disease 2019'/exp

15. '2019-ncov disease;':ab,ti OR '2019-ncov infection':ab,ti OR 'covid 19':ab,ti OR 'covid 2019':ab,ti OR 'ncov 2019 disease':ab,ti OR 'ncov 2019 infection':ab,ti OR 'novel coronavirus 2019 disease':ab,ti OR 'novel coronavirus 2019 infection':ab,ti OR 'novel coronavirus disease 2019':ab,ti OR 'novel coronavirus infection 2019':ab,ti OR 'wuhan coronavirus disease':ab,ti OR 'wuhan coronavirus infection':ab,ti

16. #14 OR #15
