## Supplement S3 for "COVID-19 among patients with hepatitis B or hepatitis C: A systematic review"

**SUPPLEMENTARY FILE S3**

The Joanna Briggs Institute critical appraisal tools (<https://joannabriggs.org/ebp/critical_appraisal_tools>) were used to assess the methodological quality of the included papers. This supplementary file presents details of quality assessment tools and scores given to each item.

**Table S1: Critical appraisal for case-report studies included in the review**

| First Author | 1 | 2 | 3 | 4 | 5 | 6 | 7 | 8 | Total score |
| --- | --- | --- | --- | --- | --- | --- | --- | --- | --- |
| Liu, B. | Yes | No | Yes | No | Yes | Yes | No | Yes | 5/8 |
| Müller, H. | Yes | Yes | Yes | No | Yes | Yes | No | Yes | 6/8 |
| Qin, J. | Yes | Yes | Yes | No | Yes | Yes | No | Yes | 6/8 |
| Zhao, J. | Yes | Yes | Yes | No | Yes | Yes | No | Yes | 6/8 |
| Hammami, MB. | Yes | Yes | Yes | Yes | Yes | Yes | No | Yes | 7/8 |
| Machado, DJDB. | Yes | Yes | Yes | Yes | Yes | Yes | No | Yes | 7/8 |
| Aldhaleei, W. A. | Yes | Yes | Yes | Yes | Yes | Yes | No | Yes | 7/8 |
| De Gottardi, A. | Yes | Yes | Yes | Yes | Yes | Yes | No | Yes | 7/8 |
| Kreivenaite, E. | Yes | Yes | Yes | Yes | No | Yes | No | Yes | 6/8 |
| Huang, JF. | Yes | Yes | Yes | Yes | Yes | Yes | No | Yes | 7/8 |

1. Were patient’s demographic characteristics clearly described?
2. Was the patient’s history clearly described and presented as a timeline?
3. Was the current clinical condition of the patient on presentation clearly described?
4. Were diagnostic tests or assessment methods and the results clearly described?
5. Was the intervention(s) or treatment procedure(s) clearly described?
6. Was the post-intervention clinical condition clearly described?
7. Were adverse events (harms) or unanticipated events identified and described?
8. Does the case report provide takeaway lessons?

**Table S2: Critical appraisal for case-series studies included in the review**

| First Author | 1 | 2 | 3 | 4 | 5 | 6 | 7 | 8 | 9 | 10 | Total Score |
| --- | --- | --- | --- | --- | --- | --- | --- | --- | --- | --- | --- |
| Loinaz, C. | Yes | Yes | Yes | Yes | Yes | Yes | Yes | No | Yes | Yes | 9/10 |
| Patrono, D. | No | Yes | Yes | No | No | Yes | No | Yes | Yes | Yes | 6/10 |
| Waisberg, DR. | Yes | Yes | Yes | Yes | Yes | Yes | Yes | Yes | No | Yes | 9/10 |
| Gao, F. | No | Yes | Yes | No | No | Yes | Yes | Yes | No | Yes | 6/10 |
| Qi, X. | Yes | Yes | Yes | No | No | Yes | Yes | Yes | No | Yes | 7/10 |
| Li, Y. | Yes | Yes | Yes | Yes | Yes | Yes | Yes | Yes | Yes | Yes | 10/10 |
| Zhong, Z. | No | Yes | Yes | No | No | Yes | Yes | Yes | No | Yes | 6/10 |
| John Hann, A. | No | Yes | Yes | No | No | Yes | No | Yes | No | Yes | 5/10 |
| Lee, BT. | Yes | Yes | Yes | Yes | Yes | Yes | Yes | Yes | Yes | Yes | 10/10 |
| Fernández‐Ruiz, M. | Yes | Yes | Yes | Yes | Yes | Yes | No | Yes | No | Yes | 8/10 |
| Kates, OS. | Yes | Yes | Yes | Yes | Yes | Yes | Yes | Yes | No | Yes | 9/10 |
| Song, S. H. | No | Yes | Yes | No | No | Yes | Yes | Yes | No | NA | 5/10 |

1. Were there clear criteria for inclusion in the case series?
2. Was the condition measured in a standard, reliable way for all participants included in the case series?
3. Were valid methods used for identification of the condition for all participants included in the case series?
4. Did the case series have consecutive inclusion of participants?
5. Did the case series have complete inclusion of participants?
6. Was there clear reporting of the demographics of the participants in the study?
7. Was there clear reporting of clinical information of the participants?
8. Were the outcomes or follow up results of cases clearly reported?
9. Was there clear reporting of the presenting site(s)/clinic(s) demographic information?
10. Was statistical analysis appropriate?

**Table S3: Critical appraisal for cross-sectional studies included in the review**

| First Author | 1 | 2 | 3 | 4 | 5 | 6 | 7 | 8 | 9 | Total Score |
| --- | --- | --- | --- | --- | --- | --- | --- | --- | --- | --- |
| Chen, L. | Yes | Yes | Yes | No | Yes | Yes | Yes | Yes | Yes | 8/9 |
| Chen, X. | Yes | Yes | Yes | Yes | Yes | Yes | Yes | Yes | Yes | 9/9 |
| Guan W-jie | Yes | Yes | Yes | No | Yes | Yes | Yes | Yes | Yes | 8/9 |
| Liu, J. | Yes | Yes | Yes | Yes | Yes | Yes | Yes | Yes | Yes | 9/9 |
| Zhang, B. | Yes | Yes | Yes | Yes | Yes | Yes | Yes | Yes | Yes | 9/9 |
| Zou, X. | Yes | Yes | Yes | Yes | Yes | Yes | Yes | Yes | Yes | 9/9 |

1. Was the sample frame appropriate to address the target population?
2. Were study participants sampled in an appropriate way?
3. Was the sample size adequate?
4. Were the study subjects and the setting described in detail?
5. Was the data analysis conducted with sufficient coverage of the identified sample?
6. Were valid methods used for the identification of the condition?
7. Was the condition measured in a standard, reliable way for all participants?
8. Was there appropriate statistical analysis?
9. Was the response rate adequate, and if not, was the low response rate managed appropriately?
